# Load modulation affects pediatric lower limb joint moments during a step-up task

**DOI:** 10.1101/2023.10.20.23296774

**Authors:** Vatsala Goyal, Keith E. Gordon, Theresa Sukal-Moulton

**Author notes:** Corresponding author Theresa Sukal-Moulton, PT, DPT, PhD Department of Physical Therapy and Human Movement Sciences Northwestern University 645 N. Michigan Ave, Ste 1100 Chicago, Illinois, USA 60611.

## Abstract

Performance in a single step has been suggested to be sensitive measure of movement quality in pediatric clinical populations. Although there is less information available in children with typical development, researchers have postulated the importance of analyzing the effect of body weight modulation on the initiation of stair ascent, especially during single limb stance where upright stability is most critical. The purpose of this study was to investigate the effect of load modulation from -20% to +15% of body weight on typical pediatric lower limb joint moments during a step-up task. Fourteen participants between 5-21 years with no known history of neurological or musculoskeletal concerns were recruited to perform multiple step-up trials. Peak extensor support and hip abduction moments were identified during the push-off and pull-up stance phases. Linear regressions were used to determine the relationship between peak moments and load. Mixed effects models were used to estimate the effect of load on hip, knee, and ankle percent contributions to peak support moments. There was a positive linear relationship between peak support moments and load in both stance phases, where these moments scaled with load. There was no relationship between peak hip abduction moments and load. While the ankle and knee were the primary contributors to the support moments, the hip contributed more than expected in the pull-up phase. Clinicians can use these results to contextualize movement differences in pediatric clinical populations including cerebral palsy and highlight potential target areas for rehabilitation for populations such as adolescent athletes.

## Introduction

Body weight modulation, including providing partial support of a person’s body weight (Celestino et al., 2014; Cherng et al., 2007; Kurz et al., 2011; Phillips et al., 2007; Provost et al., 2007) and addition of external loads (Dodd et al., 2003; McBurney et al., 2003; Simão et al., 2014), is broadly used in research and clinical practice. This method is often adopted for individuals with neurodevelopmental disorders during steady-state gait training (Celestino et al., 2014; Cherng et al., 2007; Kurz et al., 2011; Phillips et al., 2007; Provost et al., 2007; Simão et al., 2014). The evidence is less robust for the effect of body weight modulation on the initiation of stair ascent. Researchers have postulated the importance of analyzing this specific movement because the first step up requires larger lower limb joint moments compared to subsequent steps (Wang and Gillette, 2018). This analysis may be especially important for pediatric clinical populations, for which a single step up can be a sensitive measure of movement quality (Stania et al., 2017).

The handful of studies that have investigated lower limb moments of a step-up task have all been in adults, and even fewer studies have explored the effect of load modulation (Goyal et al., 2022; Wang and Gillette, 2018). It has been confirmed that substantial hip abduction moments are necessary to maintain mediolateral stability and considerable sagittal plane extensor moments are required to keep the body upright in adults (Goyal et al., 2022; Novak and Brouwer, 2011; Wang and Gillette, 2018). There is a need for a robust biomechanical model defining how these joint moments change across multiple load conditions in a typical pediatric population, especially during single limb stance where upright stability is most critical. This model would serve to contextualize movement differences in pediatric populations with neurodevelopmental disorders and highlight potential target areas for training during clinical therapy.

The purpose of this study was to investigate the effect of body weight load modulation from -20% body weight (BW) to +15% BW in 5% increments on typical pediatric lower limb joint moments during a step-up task. Based on previous literature, we hypothesized that 1) extensor support moments would incrementally increase with load during the stance phases of a step up, 2) hip abduction moments would incrementally increase with load during the stance phases of a step up, and 3) the knee and ankle joints would drive increases in extensor support moments. We also performed a secondary analysis to understand the relationship between age and leg length and the kinetic performance of a step-up task.

## Methods

### I. Participant Summary

Participants were recruited as a sample of convenience through word-of-mouth and flyers. Individuals were included in the study if they were between the ages of 5-21 years with no known history of neurological or musculoskeletal concerns that would affect their ambulation or ability to participate in the study. We sampled from a wide age range to capture step-up performance across the pediatric population. This study was approved by Northwestern University’s Institutional Review Board. Participants under the age of 18 provided verbal or written assent along with parent/guardian written consent. Participants who were 18+ years provided informed written consent themselves.

### II. Experimental Set-Up

Participants performed a series of trials that involved stepping up onto a raised platform. A 2×2 cluster of four force plates (AMTI, Watertown, MA) captured participant ground reaction forces at a frequency of 1000 Hz. To independently capture joint biomechanics from the right and left lower limbs, two 10.2-cm tall platforms were placed on two side-by-side anterior force plates. We purposefully chose a low step height in order to replicate this experiment in the future with pediatric clinical populations, who may have a difficult time completing the protocol on a higher platform (Goyal et al., 2022). A 10-camera motion capture system (Qualisys, Göteborg, Sweden) recorded participant kinematics at a frequency of 100 Hz using a modified Cleveland Clinic marker set (Kaufman et al., 2016). Markers were placed on thirty-four total landmarks of the trunk (sternum, C7 vertebrae, T10 vertebrae), pelvis (sacrum, bilateral posterior superior iliac spines), and lower extremities (bilateral greater trochanters, lateral femoral epicondyles, lateral malleoli, calcanei, second and fifth metatarsals, and thigh and shank four-marker clusters).

During step-up trials, participant body weight (BW) was modulated using the Zero-G Bodyweight Support System (Aretech LLC, Ashburn, VA) to subtract weight or a weighted vest to add weight. There were six total load conditions: three unweighted conditions of -20%, -15%, and -10% of BW and three weighted conditions of +5%, +10%, and +15% of BW. The weighted conditions were chosen based on loads common for children’s backpacks (Bryant and Bryant, 2014; Perrone et al., 2018). Available weights included ⅓, ⅔, ½, and 1-lb bars, which were distributed evenly around the vest. The unweighted conditions represented a similar range to the weighted conditions; we did not test a -5% condition due to technical limitations of the Zero-G, which requires a minimum of 10 lbs to be removed from the user.

### III. Experimental Protocol

Participants filled out the Waterloo Footedness Questionnaire – Revised (Elias et al., 1998) and completed timed single-limb stance tests to determine lower limb dominance. During the experiment, participants started in a standing position with their feet split between the two force plates posterior to the platforms (Figure 1A).

**Figure 1.**
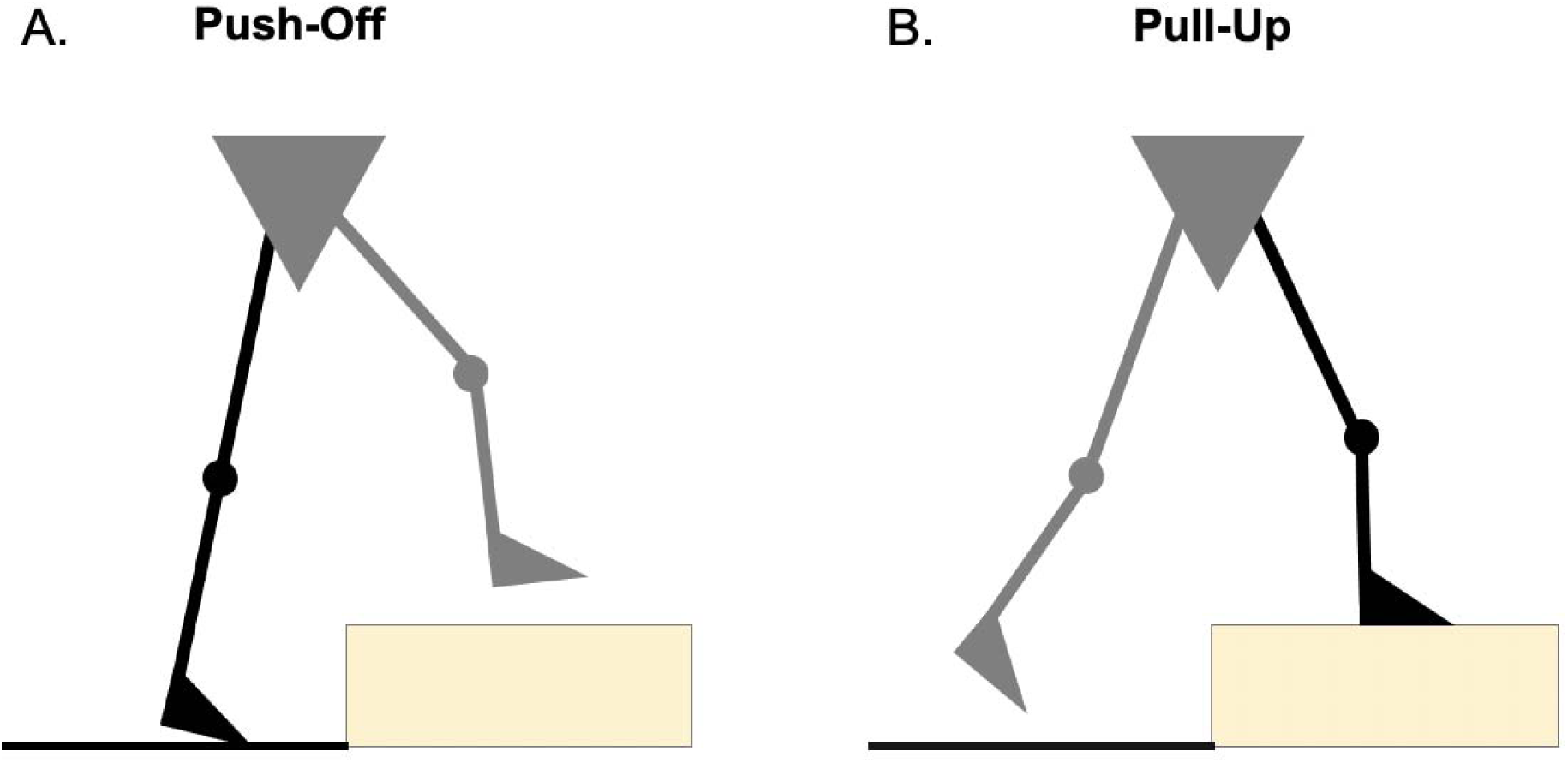
(A) The push-off stance phase of the step-up task, between leading foot lift-off and leading foot initial contact with the step. (B) The pull-up stance phase of the step-up task, between trailing foot lift-off and trailing foot initial contact with the step.

Participants were instructed to step up onto the platforms at a self-selected walking speed and, after a slight pause at the top, were instructed to step back down to the starting position. Participants completed three blocks of trials, where they were instructed to lead a series of step-ups with either their dominant foot or their non-dominant foot. The first block was a no-load condition, where participants completed 15 steps per leading foot. The second and third blocks were randomized and consisted of either weighted or unweighted conditions. Within these blocks, the load conditions were randomized and participants completed 10 steps per leading foot. To ensure participant safety and minimize fall risk, participants were connected to an overhead trolley with a harness.

### III. Data Processing and Analysis

Data were first visually inspected in Qualisys Track Manager (Qualisys, Göteborg, Sweden) to verify that the markers were appropriately labeled. Data were then imported into Visual 3D (C-Motion, Germantown, MD). Marker and ground reaction force data were interpolated to fill in small gaps and filtered using a low-pass 4th-order Butterworth filter with a 6 Hz cutoff frequency to remove high-frequency fluctuations (Goyal et al., 2022). We performed inverse dynamics calculations in Visual 3D to calculate hip, knee, and ankle joint moments in the sagittal and frontal plane. To account for the step height during these calculations, the two corresponding force plates were modified virtually to create raised force platforms. We used ground reaction force data to identify four gait events occurring during each step-up: leading limb lift-off, leading limb initial contact on the step, trailing limb lift-off, and trailing limb initial contact on the step (Goyal et al., 2022).

Further processing was done in MATLAB (MathWorks, Inc., Natick, MA). We identified hip abduction and extensor support moments (the sum of hip, knee, and ankle sagittal plane moments). All joint moment data were divided by participant body weight for comparison during statistical analyses. We plotted joint moment profiles for each individual trial during two single-limb stance phases: 1) the push-off phase, between leading foot lift-off and leading foot initial contact (Figure 1A) and 2) the pull-up phase, between trailing foot lift-off and trailing foot initial contact (Figure 1B). Any trials that were two standard deviations outside of the average stance phase length were not considered for further analysis.

### IV. Statistical Analysis

Statistical analysis was performed in Stata IC 14.1 (StataCorp LLC, College Station, TX), and significance was set at p<0.05. The push-off and pull-up stance phases were considered separately in all statistical analyses. Visual inspection of the histogram distributions of residuals was used to confirm the normality of the data. We considered the outcome measures of peak hip abduction moments, peak support moments, and individual hip, knee, and ankle percent contributions to peak support moments. The latter was calculated by dividing hip, knee, and ankle moments at the time of peak support moment by the peak support moment. For the no-load condition, we ran mixed effects models to estimate the fixed effect of the limb dominance (2 levels: dominant, non-dominant) and a random effect of participant on these outcomes. In a secondary analysis, we also ran Pearson’s correlations to evaluate the strength of the relationships between peak moments and the continuous variables of age and leg length.

For the six load conditions, peak hip abduction moments and peak support moments were averaged and normalized to their average values in the no-load condition to facilitate comparison across participants. We first ran a linear mixed effects model to determine if there were differences between the dominant and non-dominant limbs at each individual load level (interaction term). If not, the data were then combined. Linear regressions were used on these outcome measures to determine the relationship between normalized peak moments and load. For the outcome measures of individual joint percent contributions to peak support moments, we used linear mixed effects models with two fixed effects of load (−20%, -15%, -10%, 0%, +5%, +10%, and +15% of BW) and limb (dominant, non-dominant) and a random effect of participant. Bonferroni corrections were used to correct for multiple comparisons in post-hoc analyses. In another secondary analysis, we calculated the individual slopes of peak moments vs. load for each participant. We then ran Pearson’s correlations between these slopes and age and leg length.

## Results

### I. Participant Metrics

Fourteen individuals participated in the study (7 female). As the statistical analysis included repeated measures for each participant, the average effective sample size calculated was 25 participants. Thirteen participants were right-foot dominant and one was left-foot dominant. Participant metrics included a mean age of 12.8 ± 4.2 years, a mean weight of 53.2 ± 28.1 kg, and a mean height of 1.54 ± 0.19 m. Two participants were unable to complete the +15% load condition because the calculated BW percentage exceeded the available weights. Four participants were unable to complete the -10% condition and one participant was unable to complete the -15% condition because their weight was too low to meet the minimum weight removal requirements of the Zero-G.

### II. Push-Off Phase

For the no-load condition, the effect of limb dominance was not significant for any outcome measures (Figure 2A). Average peak hip abduction moments were +0.730 Nm/kg and average peak support moments were -0.791 Nm/kg. There was a large ankle plantarflexion contribution of 112% to peak support moments. This served to offset a small hip flexion contribution of 3.81% and a small knee flexion contribution of 8.62%, resulting in a net extension moment.

**Figure 2.**
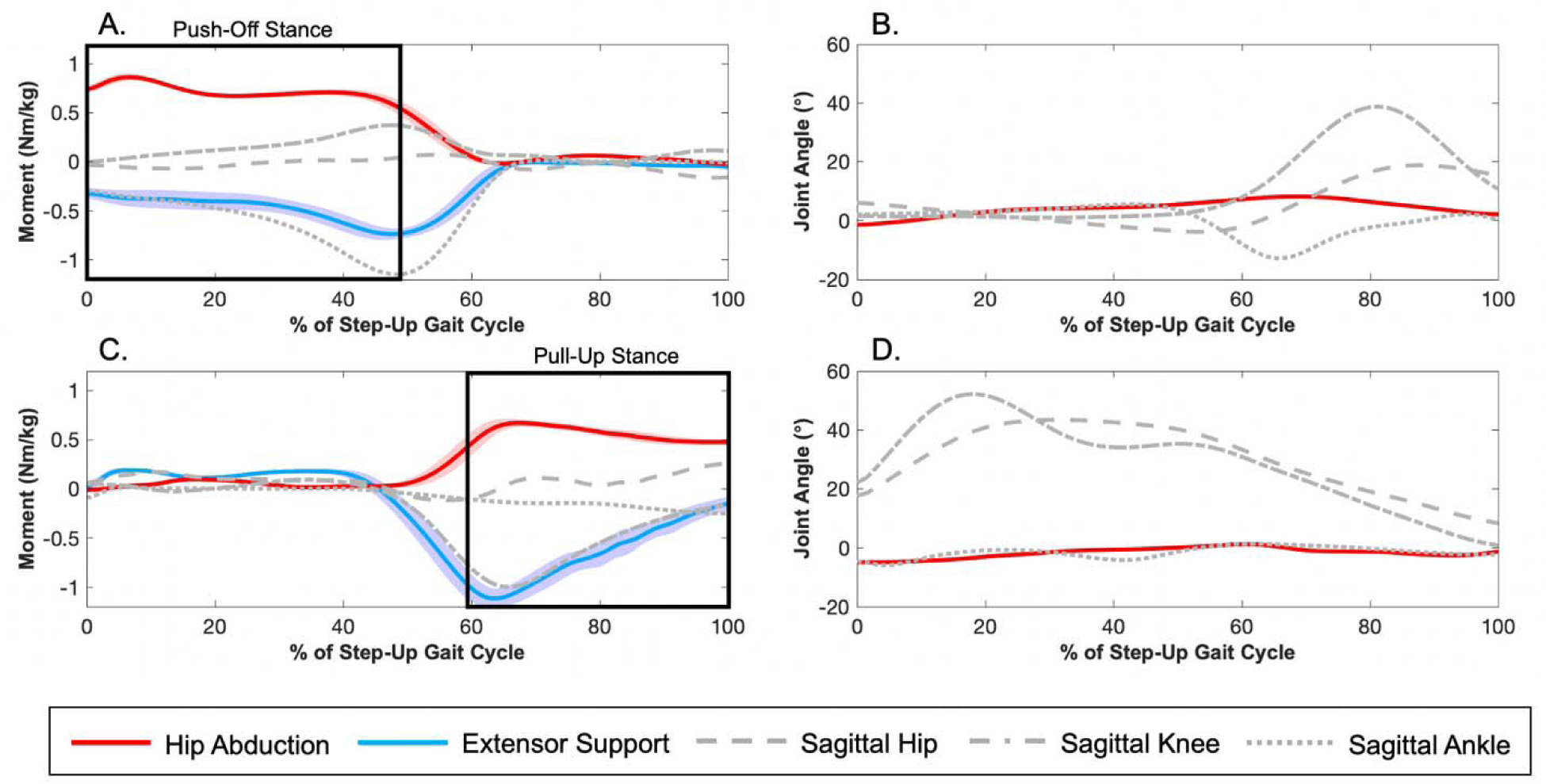
Representative kinetic (A and C) and kinematic (B and D) profiles from one participant during a no-load step up for the trailing leg (A and B) and the leading leg (C and D). On each x-axis, 0% corresponds to the start of a step-up trial at leading leg lift-off while 100% corresponds to the end of the trial at trailing leg initial contact with the step. On each y-axis, a positive magnitude indicates joint flexion/abduction while a negative magnitude indicates joint extension/adduction. Average hip abduction moments are in red. Individual lower limb sagittal plane moments are in gray, including the hip (gray dash), knee (gray dash-dot), and ankle (gray dot). The sum of these individual joint moments equals the extensor support moments shown in blue. Shaded regions represent one standard deviation. The black boxes on plots A and C indicate the push-off and pull-up stance phases, respectively.

There were no significant differences in normalized peak moments between the dominant and non-dominant limbs at each individual load level. The linear relationship between peak hip abduction moments and load was not significant in the push-off stance phase. In contrast, the linear relationship between peak support moments and load was significant (p<0.001, R^2^ = 0.278), with a coefficient of +0.817 and an intercept of +0.973 (Figure 3A). As for individual percent contributions to peak support moments, the effect of load was only significant for hip percent contributions to peak support moments (p=0.001). However, there were no significant pairwise comparisons (Figure 4A, 4C, 4E).

**Figure 3.**
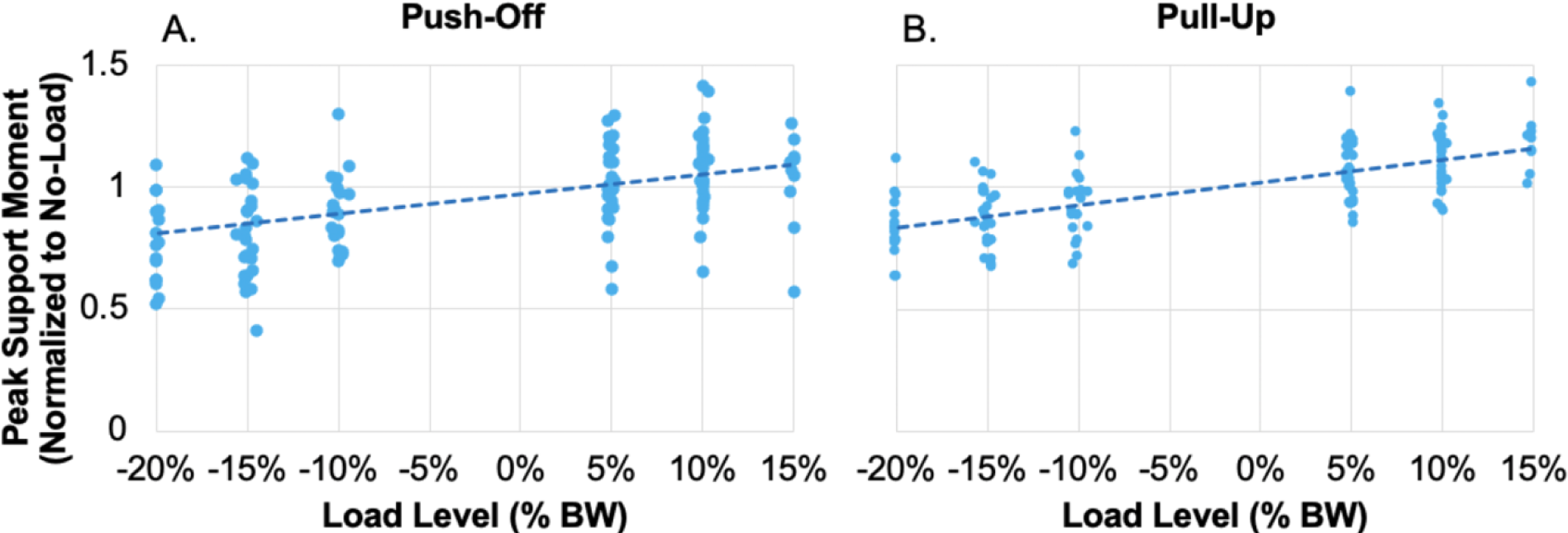
Peak support moments vs. load for the (A) push-off and (B) pull-up stance phases. All values are divided by their respective values in the no-load condition. The linear regression for both stance phases showed a significant relationship between the two variables, with y = 0.817x + 0.973 for the push-off phase (R^2^ = 0.278) and y = 0.933x + 1.02 for the pull-up phase (R^2^ = 0.498).

**Figure 4.**
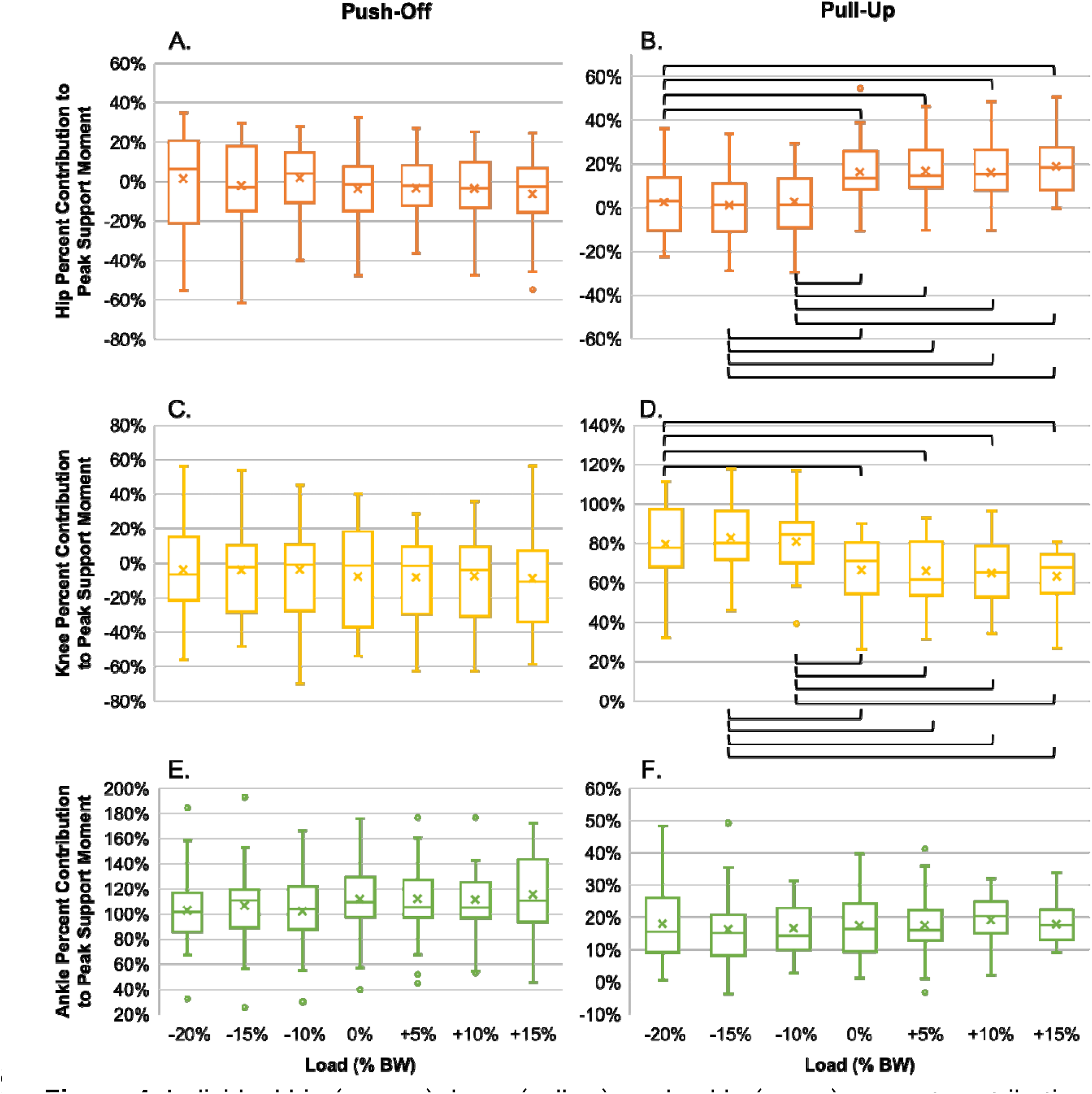
Individual hip (orange), knee (yellow), and ankle (green) percent contributions to peak extensor support moment at the time of peak support moment for all loading conditions. A negative percent contribution represents a joint moment in flexion, while a positive percent contribution represents a joint moment in extension. Significant pairwise comparisons are shown by black brackets (corrected p<0.001).

### III. Pull-Up Phase

The effect of limb dominance was not significant for any outcome measures in the no-load condition (Figure 2C). Average peak hip abduction moments were +0.607 Nm/kg and average peak support moments were -1.20 Nm/kg. Average individual joint percent contributions to peak support moments were 15.9% hip extension, 66.5% knee extension, and 17.6% ankle plantarflexion.

There were no significant differences in normalized peak moments between the dominant and non-dominant limbs at each individual load level. The linear relationship between peak hip abduction moments and load was again not significant in the pull-up stance phase. In contrast, the linear relationship between peak support moments and load was significant (p<0.001, R^2^ = 0.498), with a coefficient of +0.933 and an intercept of +1.02 (Figure 3B).

The effect of load was significant for hip and knee percent contributions to peak support moments (both p<0.001). Pairwise comparisons revealed that hip contributions were significantly larger for +0%, +5%, +10%, and +15% compared to -20%, -15%, and -10%, while knee contributions were significantly larger for -20%, -15%, and -10% compared to +0% +5%, +10%, and +15% (p<0.001 for all). Despite these opposing changes, the knee remained the primary contributor to peak support moments across all load conditions (Figure 4B, 4D, 4F). The interaction between limb dominance and load was also significant for knee percent contributions (p=0.003). There were no significant pairwise comparisons between the dominant and non-dominant limbs at each individual load level (supplementary materials S1).

### IV. Secondary Analyses: Age & Leg Length

As there was no significant effect of limb dominance on peak moments, the values for the dominant and non-dominant lower limbs were collapsed for the secondary analyses. For the no-load condition, Pearson’s correlation analysis between peak hip abduction moments and age was significant in the push-off (r = +0.830, p<0.001) and pull-up stance phases (r = +0.833, p<0.001). This analysis was also significant between peak hip abduction moments and leg length in push-off (r = +0.753, p<0.001) and pull-up (r = +0.770, p<0.001). In summary, the magnitude of peak hip abduction increased with age and with leg length (Figure 5A, 5B). Pearson’s correlation analysis was significant between peak support moments and age in the push-off (r = +0.304, p<0.001) and pull-up stance phases (r = +0.358, p<0.001). This analysis was also significant between peak support moments and leg length in push-off (r = +0.265, p<0.001) and pull-up (r = +0.217, p<0.001). The magnitude of peak support moment decreased with age and leg length (Figure 5C, 5D). For the load conditions, there were no significant correlations between the individual participant slopes of peak moments vs. load and age and leg length.

**Figure 5.**
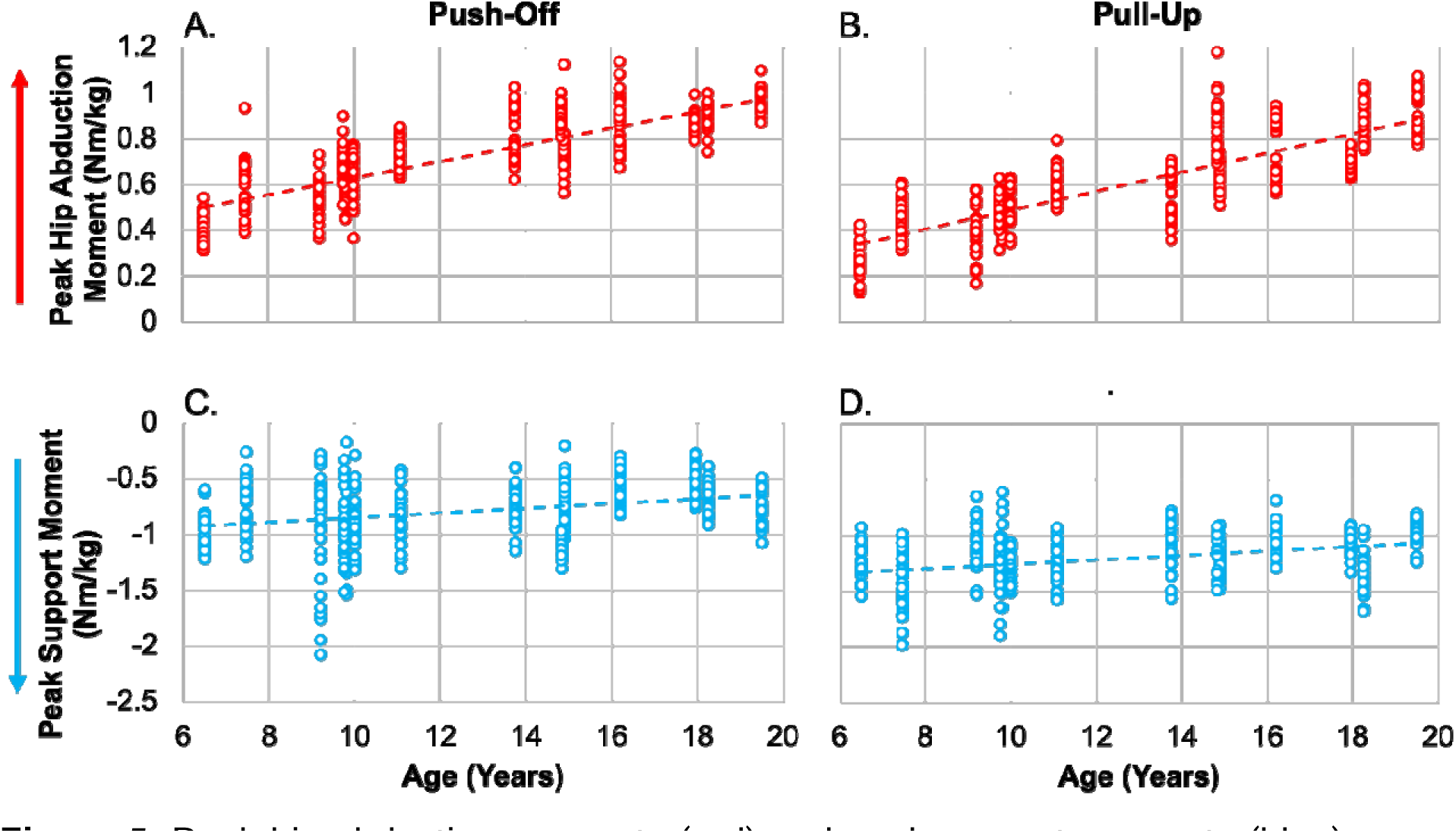
Peak hip abduction moments (red) and peak support moments (blue) vs. age for the no-load condition during the push-off and pull-up stance phases. Each point represents an individual no-load trial. All moment values are divided by participant weight, and colored arrows on the far left show the direction of increasing moment magnitude. Pearson’s correlation was significant for all relationships, with r-values of (A) +0.830, (B) +0.833, (C) +0.304, and (D) +0.358. Results indicate that the magnitude of peak hip abduction increases with age, while the magnitude of peak support moment decreases with age.

## Discussion

We investigated the effect of modulating body weight load from -20% to +15% on lower limb biomechanical strategies in pediatric individuals during a step-up task. There was a positive linear relationship between peak support moments and load in the push-off and pull-up stance phases, where these moments scaled with load. There was no relationship between peak hip abduction moments and load in either stance phase.

While the ankle and knee were the primary contributors to the support moments, the hip contributed more than expected during weighted conditions in the pull-up phase. In a secondary analysis, we also found a significant positive correlation between peak hip abduction moments and age and leg length during the no-load condition.

Peak hip abduction moments during the no-load condition of our step-up task were similar to what has been reported in stair ascent (Novak and Brouwer, 2013, 2011; Strutzenberger et al., 2011). Our hypothesis that hip abduction moments would incrementally increase with load in both stance phases of a step-up task was not supported. Contrary to our results, significant increases in hip abduction moments between no-load and +20% of BW have been quantified during the initial step up in adults (Wang and Gillette, 2018). Hip abductor muscle activations have also been shown to significantly increase between -30% of body weight and no-load during regular gait (Mun et al., 2017). One possible explanation for our findings may be that hip abduction moments generated in the no-load condition were at a magnitude that supported mediolateral stability across all loading conditions. Alternatively, participants may have altered their stepping behavior during unweighted conditions, similar to what has been seen in adults during treadmill walking (Dragunas and Gordon, 2016). It would be interesting to use these results as a comparator to outcome measures for pediatric populations with conditions such as patellofemoral pain syndrome, where the hip abductors are weak (Xie et al., 2023).

Peak support moments during the no-load condition of our step-up task were slightly lower than what has been reported in stair ascent (Novak and Brouwer, 2013, 2011; Strutzenberger et al., 2011), most likely due to the lower step height in our study. Supporting our hypothesis, peak support moments significantly increased with load incrementally in both stance phases of a step-up task. Previous studies quantifying peak vertical ground reaction forces during the push-off and pull-up phases of stair ascent within narrow ranges of load modulation are consistent with our results (Bannwart et al., 2019; Hong and Li, 2005). This experimental paradigm may be beneficial for gait rehabilitation approaches targeting the lower limb extensors, particularly for individuals with cerebral palsy who have weak lower limb extensors (Wiley and Damiano, 1998). Hip, knee, and ankle percent contributions to peak support moments can reveal which joints are driving these changes in response to load.

The effect of load modulation on individual sagittal plane moments were expected in the push-off phase and unexpected in the pull-up phase. Similar to stair ascent, ankle plantarflexion moments contributed the most to extensor support moments in the push-off phase of a no-load step-up task (McFadyen and Winter, 1988; Nadeau et al., 2003; Novak and Brouwer, 2011; Riener et al., 2002). Our results also suggest that ankle moments scaled with peak support moments to remain the primary contributor in the push-off phase across all load conditions. Indeed, the ankle has been shown to be the most responsive joint to changes in body weight support (Goldberg and Stanhope, 2013). In contrast, the knee is primarily responsible for vertical progression of the body during stair negotiation (Costigan et al., 2002; McFadyen and Winter, 1988; Nadeau et al., 2003; Novak and Brouwer, 2011; Riener et al., 2002). This was observed in our calculated knee extension moments during the pull-up phase of a no-load step-up task. Though knee moments remained the primary contributor to peak support moments in this phase across all load conditions, these moments decreased when body weight support was removed while hip extension moments proportionally increased. This unexpected strategy may have been used to prevent overloading of the knee joint, which can operate as high as 72% of maximum capacity during regular stair climbing in young adults (Reeves et al., 2009). The hip and knee work in tandem during pull-up (Riener et al., 2002), which may explain why the redistribution of extensor moments in the weighted block is towards the hip. Using a step-up task with body weight support may be useful for clinical interventions focused on strength and control of the knee and ankle joints in pediatric populations with weaker distal joints, such as individuals with cerebral palsy (Fowler et al., 2010; Wiley and Damiano, 1998). Alternatively, adding external loads to a step-up task may be a worthwhile approach to train the hip joint in pediatric populations such as adolescent athletes.

The magnitude of peak hip abduction moments increased with age and with leg length in both stance phases. It is possible that step width may be driving this positive correlation. Three lines of evidence support this suggestion: 1) step width increases with age in children (Gill et al., 2016), 2) wider step widths increase the mediolateral moment arm (Henderson et al., 2011), and 3) the mediolateral moment arm is positively correlated with peak hip abduction moments (Vistamehr and Neptune, 2021). In contrast, the magnitude of peak support moments slightly decreased with age and with leg length in both stance phases, indicating that younger children with shorter limbs used more relative extension moments to complete a step-up task. It’s possible that younger children are still exploring how to optimize gait and therefore generating more extension than necessary to complete the task (Frost et al., 1997). For all participants, the step height was approximately 10-19% of their leg length; children with shorter lower limbs may have generated larger extension moments to complete a step up at a relatively larger step height. Age and leg length did not play a factor in the slopes of peak joint moments vs. load, suggesting that all participants had similar strategies when responding to different loads. It may be that the biomechanics of responding to load are developed at a young age and maintained through adolescence.

In summary, our study developed a model of the effect of load modulation from - 20% to +15% of BW on typical pediatric lower limb joint moments during a step-up task. Limitations of the study include the resolution of load, the self-selected speed of each participant, and the placement of some reflective markers on tight-fitting clothes rather than directly on skin. One future direction is comparing this model to pediatric clinical populations to describe the possible effects of atypical development or injuries and the potential impact from interventions. Another future direction is translating the experiment to a more natural environment, which includes testing participants without a harness and varying the step height.

## Data Availability

All data produced in the present study are available upon reasonable request to the authors.

## Acknowledgements

The authors would like to sincerely thank our research participants and their families for their efforts and enthusiasm. We would also like to thank Tara Cornwell and Keri Han for their help in experimental design and data analysis. This work was supported by the American Heart Association [grant number 908832] to VG, National Institutes of Health [grant number T32 HD007418] to VG, and a Northwestern University Department of Biomedical Engineering Summer Undergraduate Research Award. The funding sources had no role in the study design; in the collection, analysis, and interpretation of data; in the writing of manuscript; and in the decision to submit the manuscript for publication.

## References

1. Bannwart, M., Rohland, E., Easthope, C.A., Rauter, G., Bolliger, M., 2019. Robotic body weight support enables safe stair negotiation in compliance with basic locomotor principles. J. Neuroeng. Rehabil. 16. 10.1186/s12984-019-0631-8

2. Bryant, B.P., Bryant, J.B., 2014. Relative Weights of the Backpacks of Elementary-Aged Children. J. Sch. Nurs. 30, 19–23. 10.1177/1059840513495417

3. Celestino, M.L., Gama, G.L., Longuinho, G.S.C., Fugita, M., Barela, A.M.F., 2014. Influence of body weight unloading and support surface during walking of children with cerebral palsy. Fisioter. em Mov. 27, 591–599. 10.1590/0103-5150.027.004.ao11

4. Cherng, R.J., Liu, C.F., Lau, T.W., Hong, R. Bin, 2007. Effect of treadmill training with body weight support on gait and gross motor function in children with spastic cerebral palsy. Am. J. Phys. Med. Rehabil. 86, 548–555. 10.1097/PHM.0b013e31806dc302

5. Costigan, P.A., Deluzio, K.J., Wyss, U.P., 2002. Knee and hip kinetics during normal stair climbing. Gait Posture 16, 31–37. 10.1016/S0966-6362(01)00201-6

6. Dodd, K.J., Taylor, N.F., Graham, H.K., 2003. A randomized clinical trial of strength training in young people with cerebral palsy. Dev. Med. Child Neurol. 45, 652–657. 10.1017/S0012162203001221

7. Dragunas, A.C., Gordon, K.E., 2016. Body weight support impacts lateral stability during treadmill walking. J. Biomech. 49, 2662–2668. 10.1016/j.jbiomech.2016.05.026

8. Elias, L.J., Bryden, M.P., Bulman-Fleming, M.B., 1998. Footedness is a better predictor than is handedness of emotional lateralization. Neuropsychologia 36, 37–43. 10.1016/S0028-3932(97)00107-3

9. Fowler, E.G., Staudt, L.A., Greenberg, M.B., 2010. Lower-extremity selective voluntary motor control in patients with spastic cerebral palsy: Increased distal motor impairment. Dev. Med. Child Neurol. 52, 264–269. 10.1111/j.1469-8749.2009.03586.x

10. Frost, G., Dowling, J., Dyson, K., Bar-Or, O., 1997. Cocontraction in three age groups of children during treadmill locomotion. J. Electromyogr. Kinesiol. 7, 179–186. 10.1016/S1050-6411(97)84626-3

11. Gill, S. V., Keimig, S., Kelty-Stephen, D., Hung, Y.C., DeSilva, J.M., 2016. The relationship between foot arch measurements and walking parameters in children. BMC Pediatr. 16, 2. 10.1186/s12887-016-0554-5

12. Goldberg, S.J., Stanhope, S.J., 2013. Sensitivity of joint moments to changes in walking speed and body-weight-support are interdependent and vary across joints. J. Biomech. 46, 1176–1183.

13. Goyal, V., Dragunas, A., Askew, R.L., Sukal-Moulton, T., López-Rosado, R., 2022. Altered biomechanical strategies of the paretic hip and knee joints during a step-up task. Top. Stroke Rehabil. 1–9. 10.1080/10749357.2021.2008596

14. Henderson, E.R., Marulanda, G.A., Cheong, D., Temple, H.T., Letson, G.D., 2011. Hip abductor moment arm - a mathematical analysis for proximal femoral replacement. J. Orthop. Surg. Res. 6, 6. 10.1186/1749-799X-6-6

15. Hong, Y., Li, J.X., 2005. Influence of load and carrying methods on gait phase and ground reactions in children’s stair walking. Gait Posture 22, 63–68. 10.1016/j.gaitpost.2004.07.001

16. Kaufman, K., Miller, E., Kingsbury, T., Russell Esposito, E., Wolf, E., Wilken, J., Wyatt, M., 2016. Reliability of 3D gait data across multiple laboratories. Gait Posture 49, 375–381. 10.1016/j.gaitpost.2016.07.075

17. Kurz, M.J., Stuberg, W., Dejong, S.L., 2011. Body weight supported treadmill training improves the regularity of the stepping kinematics in children with cerebral palsy. Dev. Neurorehabil. 14, 87–93. 10.3109/17518423.2011.552459

18. McBurney, H., Taylor, N.F., Dodd, K.J., Graham, H.K., 2003. A qualitative analysis of the benefits of strength training for young people with cerebral palsy. Dev. Med. Child Neurol. 45, 658–663. 10.1017/S0012162203001233

19. McFadyen, B.J., Winter, D.A., 1988. An integrated biomechanical analysis of normal stair ascent and descent. J. Biomech. 21, 733–744. 10.1016/0021-9290(88)90282-5

20. Mun, K.R., Lim, S. Bin, Guo, Z., Yu, H., 2017. Biomechanical effects of body weight support with a novel robotic walker for over-ground gait rehabilitation. Med. Biol. Eng. Comput. 55, 315–326. 10.1007/s11517-016-1515-8

21. Nadeau, S., McFadyen, B.J., Malouin, F., 2003. Frontal and sagittal plane analyses of the stair climbing task in healthy adults aged over 40 years: What are the challenges compared to level walking? Clin. Biomech. 18, 950–959. 10.1016/S0268-0033(03)00179-7

22. Novak, A.C., Brouwer, B., 2013. Kinematic and kinetic evaluation of the stance phase of stair ambulation in persons with stroke and healthy adults: A pilot study. J. Appl. Biomech. 29, 443–452. 10.1123/jab.29.4.443

23. Novak, A.C., Brouwer, B., 2011. Sagittal and frontal lower limb joint moments during stair ascent and descent in young and older adults. Gait Posture 33, 54–60. 10.1016/j.gaitpost.2010.09.024

24. Perrone, M., Orr, R., Hing, W., Milne, N., Pope, R., 2018. The impact of backpack loads on school children: A critical narrative review. Int. J. Environ. Res. Public Health 15, 1–25. 10.3390/ijerph15112529

25. Phillips, J.P., Sullivan, K.J., Burtner, P.A., Caprihan, A., Provost, B., Bernitsky-beddingfield, A., 2007. Ankle dorsiflexion fMRI in children with cerebral palsy undergoing intensive body-weight-supported treadmill training: A pilot study. Dev. Med. Child Neurol. 49, 39–44. 10.1017/S0012162207000102.x

26. Provost, B., Dieruf, K., Burtner, P.A., Phillips, J.P., Bernitsky-Beddingfield, A., Sullivan, K.J., Bowen, C.A., Toser, L., 2007. Endurance and gait in children with cerebral palsy after intensive body weight-supported treadmill training. Pediatr. Phys. Ther. 19, 2–10. 10.1097/01.pep.0000249418.25913.a3

27. Reeves, N.D., Spanjaard, M., Mohagheghi, A.A., Baltzopoulos, V., Maganaris, C.N., 2009. Older adults employ alternative strategies to operate within their maximum capabilities when ascending stairs. J. Electromyogr. Kinesiol. 19, e57–e68. 10.1016/j.jelekin.2007.09.009

28. Riener, R., Rabuffetti, M., Frigo, C., 2002. Stair ascent and descent at different inclinations. Gait Posture 15, 32–44.

29. Simão, C.R., Galvão, É.R.V.P., Fonseca, D.O. da S., Bezerra, D.A., Andrade, A.C. de, Lindquist, A.R.R., 2014. Effects of adding load to the gait of children with cerebral palsy: a three-case report. Fisioter. e Pesqui. 21, 67–73. 10.1590/1809-2950/470210114

30. Stania, M., Sarat-Spek, A., Blacha, T., Kazek, B., Slomka, K.J., Emich-Widera, E., Juras, G., 2017. Step-initiation deficits in children with faulty posture diagnosed with neurodevelopmental disorders during infancy. Front. Pediatr. 5, 1–7. 10.3389/fped.2017.00239

31. Strutzenberger, G., Richter, A., Schneider, M., Mündermann, A., Schwameder, H., 2011. Effects of obesity on the biomechanics of stair-walking in children. Gait Posture 34, 119–125. 10.1016/j.gaitpost.2011.03.025

32. Vistamehr, A., Neptune, R.R., 2021. Differences in balance control between healthy younger and older adults during steady-state walking. J. Biomech. 128, 110717. 10.1016/j.jbiomech.2021.110717

33. Wang, J., Gillette, J.C., 2018. Carrying asymmetric loads during stair negotiation: Loaded limb stance vs. unloaded limb stance. Gait Posture 64, 213–219. 10.1016/j.gaitpost.2018.06.113

34. Wiley, M.E., Damiano, D.L., 1998. Lower-Extremity strength profiles in spastic cerebral palsy. Dev. Med. Child Neurol. 40, 100–107. 10.1111/j.1469-8749.1998.tb15369.x

35. Xie, P., István, B., Liang, M., 2023. The Relationship between Patellofemoral Pain Syndrome and Hip Biomechanics: A Systematic Review with Meta-Analysis. Healthc. 11. 10.3390/healthcare11010099

