## Supplementary Materials S1 for "Load modulation affects pediatric lower limb joint moments during a step-up task"

**Supplementary Material**

| **Interaction between [Limb Load]** | **Contrast** | **Standard Error** | **Z-Score** |
| --- | --- | --- | --- |
| [D +5%] vs [D -20%] | -0.101 | 0.023 | -4.35 |
| [D +15%] vs [D -20%] | -0.123 | 0.022 | -5.59 |
| [D +5%] vs [D -15%] | -0.116 | 0.018 | -6.62 |
| [D +10%] vs [D -15%] | -0.123 | 0.029 | -4.19 |
| [D +15%] vs [D -15%] | -0.138 | 0.021 | -6.56 |
| [ND +5%] vs [D -15%] | -0.159 | 0.038 | -4.22 |
| [ND +10%] vs [D -15%] | -0.172 | 0.038 | -4.47 |
| [ND +15%] vs [D -15%] | -0.161 | 0.031 | -5.14 |
| [ND +5%] vs [D -10%] | -0.120 | 0.025 | -4.86 |
| [D +15%] vs [D -10%] | -0.141 | 0.029 | -4.85 |
| [ND +15%] vs [D -10%] | -0.165 | 0.036 | -4.53 |
| [ND 0%] vs [ND -20%] | -0.166 | 0.038 | -4.32 |
| [ND +15%] vs [ND -20%] | -0.176 | 0.039 | -4.55 |
| [ND 0%] vs [ND -15%] | -0.184 | 0.027 | -6.71 |
| [ND +5%] vs [ND -15%] | -0.192 | 0.038 | -5.02 |
| [ND +10%] vs [ND -15%] | -0.205 | 0.042 | -4.85 |
| [ND +15%] vs [ND -15%] | -0.195 | 0.035 | -5.55 |
| [ND 0%] vs [ND -10%] | -0.119 | 0.028 | -4.33 |
| [ND +15%] vs [ND -10%] | -0.129 | 0.032 | -4.09 |

**S1.** Significant pairwise comparisons for the interaction term between dominance (2 levels: Dominant [D] and Non-Dominant [ND]) and load (7 levels: -20%, -15%, -10%, 0%, +5%, +10%, +15%), which was significant for the outcome measure of knee percent contributions to peak support moment during the pull-up stance phase of a step-up task. There were no significant comparisons between the dominant and non-dominant limbs at each individual load level.
